# Epidemiological dynamics of Influenza B virus across multiple seasons in Kenya and Uganda inferred from sequence data, 2010-2021

**DOI:** 10.1101/2025.03.06.25323488

**Authors:** Brenda K. Nyarango, D. Collins Owuor, Everlyne Isoe, Martin Mutunga, Robinson Cheruiyot, Esther N. Katama, Timothy Makori, Arnold Lambisia, Joyce Nyiro, George Githinji, Sarah Kiguli, Peter Olupot-Olupot, Kathryn Maitland, Charles N. Agoti

## Abstract

Influenza B virus (IBV) genomic surveillance occurs unevenly across the globe, obscuring its epidemiology. We analysed 83 near complete IBV genomes collected between 2010 and 2022 in Kenya and Uganda. Alternating IBV lineage predominance and clade turnover was observed consistent with global patterns. No B/Yamagata strains were detected at the study sites after 2019. Multiple B/Victoria clade/subclades (V1A, V1A.3, V1A.3a, V1A.3a.2) and B/Yamagata clades (Y2 and Y3) were identified with no inter-lineage reassortants observed. Over time, the clades/subclades appeared to diversify through the accumulation of amino acid changes along the hemagglutinin (HA) segment backbone, especially within the known antigenic sites. Local outbreak strains appeared to be introduced from both within and outside Africa. The congruence of local and global strains in circulating lineages and amino acid changes suggests potentially similar effectiveness of vaccines recommended for the Northern and Southern Hemispheres in East Africa.

## Introduction

Three influenza types (A, B and C) can cause human infection and disease. Influenza B viruses (IBV) account for about 30% of the total influenza disease burden, which is estimated at ∼ 290,000 to 650,000 deaths and 3-5 million severe cases annually (WHO, 2019; Caini et al., 2019; Puzelli et al., 2019; Touré et al., 2022). Previous studies in Kenya and Uganda, indicated significant influenza disease burden throughout childhood (0-14 years) and among vulnerable populations e.g., pregnant women and immunocompromised individuals (Wabwire-Mangen et al., 2016; Cummings et al., 2016; Emukule, Otiato, et al., 2019; Nyiro et al., 2018; Otieno et al., 2021). Influenza disease and perhaps transmission is predominantly seasonal in temperate climatic regions (Caini et al., 2016). Contrastingly, year-round influenza activity is observed in tropical and subtropical regions although the types (or subtypes) within may exhibit seasonal occurrence (Belizaire et al., 2022). Understanding of the origin(s), phylogenetic relatedness and evolution of circulating influenza strains in tropical populations, including sub-Saharan Africa remains limited (Langat et al., 2017).

Two IBV lineages (B/Yamagata and B/Victoria) have co-circulated with variable predominance since early 1980s (Rota et al., 1990). Strains within these lineages are classified into clades and subclades following hemagglutinin (HA) sequence phylogenetic analysis (ECDC, 2023). A brief decline in influenza activity was observed between 2020 to 2021 during the coronavirus disease 2019 (COVID-19) pandemic caused by severe acute respiratory syndrome coronavirus 2 (SARS-CoV-2) (Young et al., 2020; Olsen et al., 2020; Woolbert et al., 2022). This has been hypothesized to have been caused by the non-Pharmaceutical interventions (NPIs) that were put in place to slow the spread of the epidemic e.g., lockdowns, social distancing and improved personal hygiene measures (Huang et al., 2021; Koutsakos et al., 2021). Remarkably, no B/Yamagata cases have been identified and confirmed by sequencing since March of 2020, leading to reports of potential extinction of this lineage (Paget et al., 2022; Koutsakos et al., 2021). Previous studies observed that there is constant turnover of genetically distinct clades in both lineages over time (Langat et al., 2017; Virk et al., 2020; Vijaykrishna et al., 2015). This turnover has been attributed to factors such as population immunity, vaccination and stochastic processes that underlie strain introduction, transmission and extinction (Nyasimi et al., 2020; Wang et al., 2008).

Influenza vaccines are reformulated periodically (∼1-2 years) based on circulating strains (WHO, 2023). Vaccines used in tropical and subtropical regions are guided by the Northern or Southern Hemisphere recommendations due to limited local strain surveillance (WHO, 2023). Trivalent vaccine formulations (TIV) having one IBV lineage plus a H1N1 and H3N2 strain and quadrivalent vaccines (QIV) (Kaplya-Bubenets, 2018) having both IBV lineages plus a H1N1 and H3N2 strain are available (Grohskopf et al., 2022). Mismatch between circulating and recommended strains in the vaccines has been periodically observed (Awadalla et al., 2023; Puzelli et al., 2019). There is an ongoing current debate whether B/Yamagata strains should continue to be included in annual influenza vaccines regimens (Koul,2024; Maclntyre et al., 2025). Notably, Kenya and Uganda do not have a national influenza policy yet. However, the Kenya National Immunisation Technical Advisory Group (KENITAG) in 2016 provisionally recommended vaccination of children aged 6-23 months (Dawa et al., 2019; Emukule et al., 2019).

Influenza surveillance and genomic analysis is instrumental in revealing circulating subtypes, clades and subclades, and processes such as antigenic drift, reassortment and glycosylation that potentially impact their epidemiological success (Nyasimi et al., 2020). This information is invaluable to assess predominant strains in circulation immune escape capacity of emergent strains and predict potential vaccine effectiveness of recommended vaccines during resurgence of IBV in communities. In this study, we investigated the epidemiological dynamics of the circulating IBV strains over eight years in Kenya and three years in Uganda, their global context and evolution using genomic sequence data.

## Materials and methods

### Study area and design

The specimens analysed in this study were collected during three studies: (a) the Health Facility (HF) surveillance study which is an ongoing outpatient acute respiratory illness (ARI) surveillance study within Kilifi Health and Demographic Surveillance System (KHDSS) that has been running since December 2020 (Kenya), (b) the Kilifi County Hospital (KCH) paediatric pneumonia admissions study which has been ongoing since 2007 (Kenya) and, (c) the Children’s Oxygen Administration Strategies Trial (COAST) study (Maitland & Mpoya, 2016) and COAST-Nutrition trial conducted between 2017 and 2020 (Uganda and Kenya).

In the HF surveillance, ∼15 samples per week per facility are collected from patients across all age groups presenting with ARI at five outpatient facilities, namely, KCH, Mavueni, Pingilikani, Mtondia and Matsangoni (**Figure 1a**). The inpatient KCH samples are collected from all eligible consented hospitalised children aged ≤5 years with syndromic severe or very severe pneumonia (Onyango et al., 2012). The COAST study samples were obtained from children aged 28 days to 12 years admitted to hospital with respiratory distress complicated by hypoxia at the Mulago National Referral Hospital, Jinja, Mbale, and Soroti Regional Referral Hospitals in Uganda and the KCH and Coast Province General Hospital (CPGH) in Kenya (**Figure 1b**). The participant inclusion criteria for these studies is described in detail in previous reports (Onyango et al., 2012; Nokes et al., 2009; Nyiro et al., 2018; Maitland et al., 2021).

**Figure 1.**
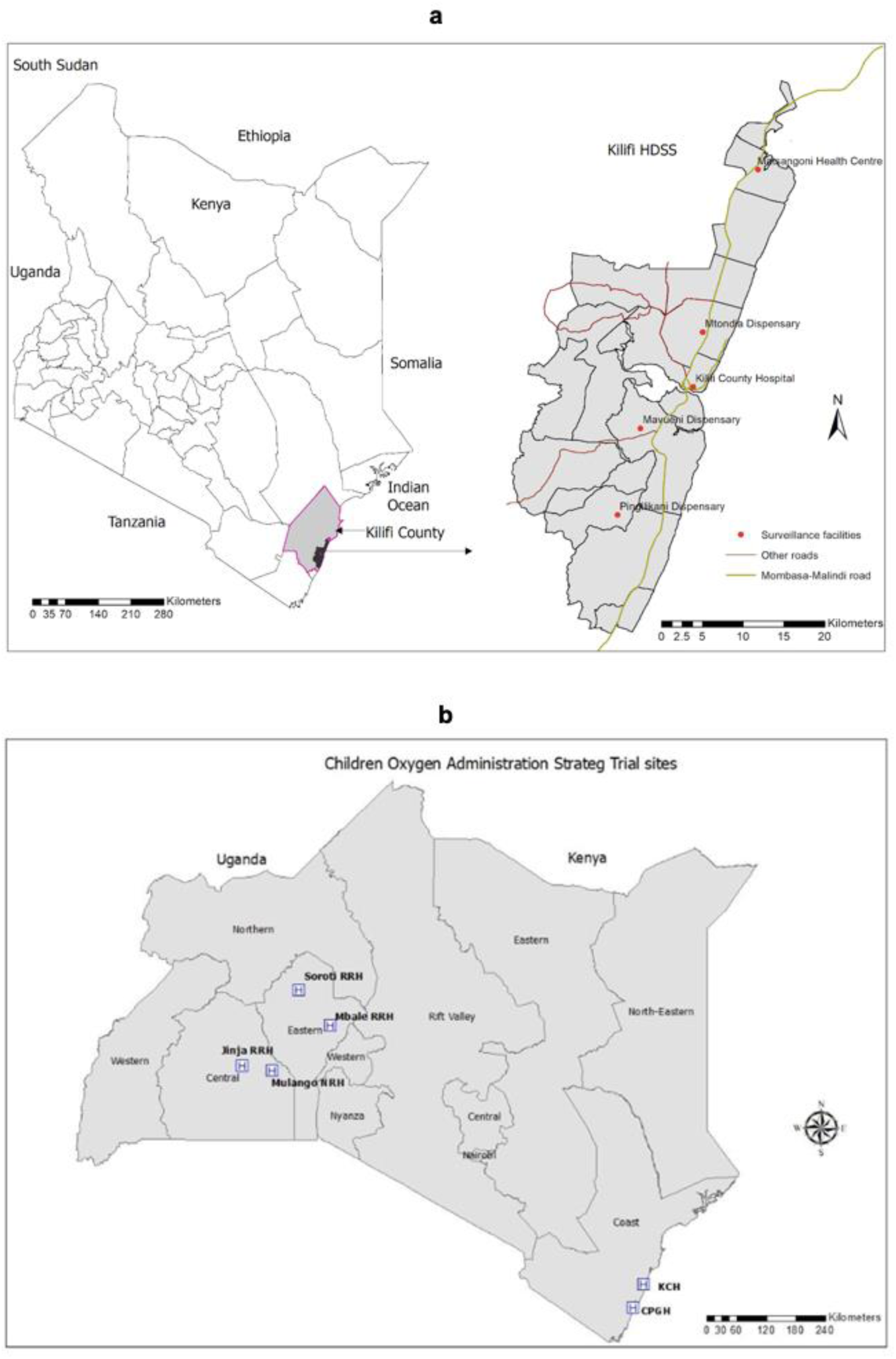
Geographical location of study sites in Kenya and Uganda. **a**, Map showing the Kilifi Health and Demographic Surveillance System area, the location of the Health Facilities that participated in the study and Kilifi County Hospital **b**, Map showing the location of Children’s Oxygen Administration Strategies Trial (COAST) hospitals in Uganda and Kenya.

### Ethical approval

Ethical approvals for the study protocols and procedures were obtained from the Kenya Medical Research Institute (KEMRI) Scientific and Ethics Review Unit (SERU), Nairobi, Kenya under study protocol numbers 3433 (KCH), 3103 (KHDSS) (KEMRI/SERU/CGMRC-C/0053/3300 and amendment C 215/4109), Makerere University REC (2016-030 and the amendment 2020-155) in Uganda and Imperial College London Research Ethics Committee (15IC3100). Participants or their parents/caregivers (if under 18 years of age) provided written informed consent to take part in the study.

### Sample collection and IBV laboratory detection

Nasopharyngeal (NP) and oropharyngeal (OP) swabs were transferred to viral transport media (VTM) and transported to KEMRI-Wellcome Trust Research Programme (KWTRP) in cool boxes with ice packs. The samples were stored at-80°C before retrieval for diagnostics and sequencing. A triplex real-time reverse transcription polymerase chain reaction (RT-PCR) diagnostic assay was used to screen samples for influenza A, B, and C (Onyango et al., 2012).

### RNA extraction and multi segment PCR

Viral RNA was extracted from 151 IBV positive samples using the QIAamp Viral RNA Mini Kit according to the manufacturer’s instructions (Qiagen, Hilden, Germany). The universal IBV-GA2 primer set (Zhou et al., 2014) was used to perform reverse transcription and amplification of the 8 IBV segments in one reaction using the SuperScriptIII One-Step RT-PCR system kit with Platinum *Taq* DNA High Fidelity polymerase (Invitrogen, Carlsbard, CA, USA). Multi-segment PCR products were stained with the RedSafe Nucleic Acid Staining solution (iNtRON Biotechnology Inc., Seoul, South Korea) on a 1.5% agarose gel and amplicons were visualised through a UV trans-illuminator.

### Next generation sequencing and assembly

PCR amplicons were purified using × 1 Agencourt AMPure XP beads to remove remaining PCR reagents (Beckman Coulter Inc., Brea, CA, USA) then quantified using Quant-iT dsDNA High-Sensitivity Assay (Invitrogen). Samples with > 40 ng/μl were classified as grade 1 and underwent normalisation by dilution with 5:7 μl RNase free water to sample ratio. The Oxford Nanopore Technologies (ONT) ligation sequencing kit and the ONT Native Barcoding Expansion kit EXP-NBD196 (Oxford Nanopore Technologies, Oxford, UK) were used to generate barcoded libraries according to the optimised ARTIC protocol (Lambisia et al., 2022). Pooled libraries were sequenced on a FLO-MIN106 R9.4.1 flow cell following the manufacturer’s instructions (ONT). Sequence base-calling and demultiplexing of FAST5 files was performed by the ONT Guppy v6.3.9 software then ONT adapters were trimmed by porechop v0.2.4. Reference-based assembly was carried out by Iterative Refinement Meta-Assembler (IRMA) v1.0.3 FLU-MinION module, suited for the highly variable nature of influenza virus due to the iterative refinement method for the reference sequences (Shepard et al., 2016). IRMA default settings: median read Q-score filter of 30, the minimum read length of 125, the frequency threshold for insertion and deletion refinement of 0.25 and 0.60, respectively; Smith–Waterman mismatch penalty of 5; and gap opening penalty of 10 were applied for this dataset (Shepard et al. 2016).

### Global and reference dataset collation

A reference dataset comprising 9 IBV clade and subclade representative sequences (**Table 1**) downloaded from Global Initiative for Sharing All Influenza Data (GISAID) was used to assign clades to the sequences. A second global dataset of all complete IBV sequences in the database from 1^st^ January 1987 to 16^th^ May 2023 of the two lineages was obtained separately. The dataset was cleaned by removing duplicated sequences, missing collection dates and Influenza A sequences (supplementary file 1). The cleaned dataset was subset using a criterion randomly selecting up to 10 sequences per month, year and continent. In addition, 108 sequences generated in 2016 from the KHDSS study surveillance in Kilifi with GISAID accession numbers EPI_ISL_336258 and EPI_ISL_336282–EPI_ISL_336395 were downloaded from the database (Nyasimi et al., 2020).

**Table 1:**
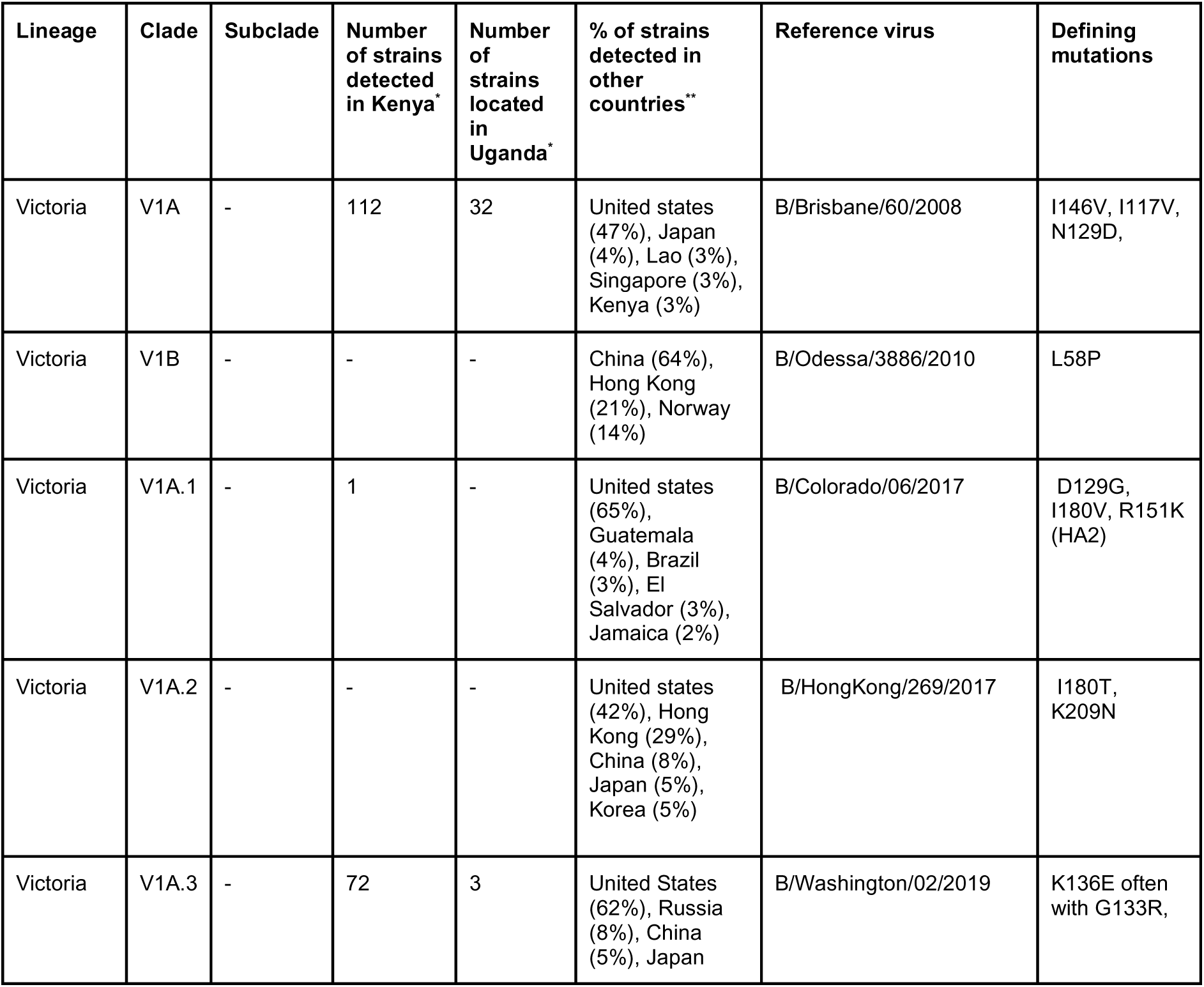
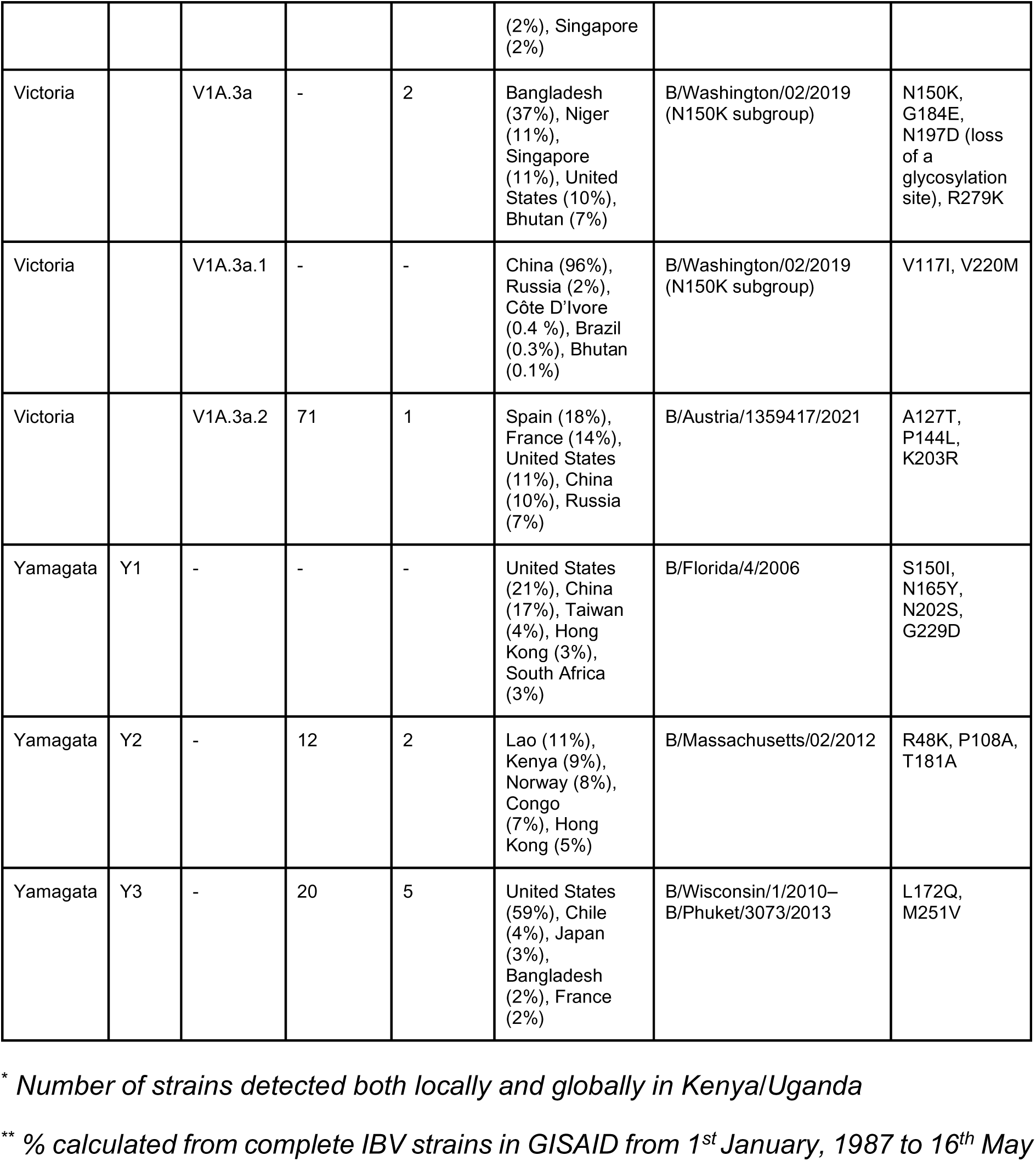
IBV lineage, clade and subclade characterisation according to ECDC classification.

### Phylogenetic analysis

Near complete (> 70%) assembled segments were aligned with the reference and global datasets by MAFFT v7.515 (Katoh & Standley, 2013). The aligned sequences were manually curated by removing leading and trailing gaps and visualised in Aliview v1.28 (Larsson, 2014). Per segment maximum likelihood phylogenies were inferred using IQTREE v2.2.6 with a clustering reliability evaluated by using 1000 bootstrap iterations (Nguyen et al., 2014). The best fitting nucleotide (nt) substitution model was determined by Modelfinder (Kalyaanamoorthy et al., 2017) and resulting ML phylogenies visualised with ggtree R package v3.6.2 (Yu, 2022). Clade and lineage assignment was done according to ECDC guidelines (ECDC, 2023) as described in (**Table 1**). Per segment phylogeny comparisons with HA were done to detect reassortants in dendextend v1.17.1 R package (Galili, 2015). Whole genomes were concatenated by Sequence matrix v1.7.6 and analysed to statistically detect reassortant strains using the Graph-incompatibility-based Assortment Finder (GiRaF) tool v1.01 (Nagarajan & Kingsford, 2010).

### Mutation and prediction of N-glycosylation sites analysis

Nextclade tool v2.14.1 (Hadfield et al., 2018) was used to identify amino acid changes along the HA backbone of the Kilifi, Uganda and global strains. These changes were plotted using the trackViewer R package v1.36.2. Potential N-linked glycosylation sites on the HA were identified using NetNGlyc v1.0 tool (http://www.cbs.dtu.dk/services/NetNGlyc/) with threshold of 0.5 applied to signify glycosylated sites.

### Global context and import/export analysis

This analysis was carried out on the HA segments of each lineage separately. Maximum likelihood phylogenies of aligned global sequences and HA segments of Kilifi and Uganda genomes were generated using IQTREE v2.2.6 (Nguyen et al., 2014). We examined the tree structures that emerged and assessed them for temporal molecular clock signals using the clock feature of TreeTime v0.2.4 (Sagulenko et al., 2017). Time-scaled phylogenies were then inferred by TreeTime implementing a relaxed clock model based on the temporal signal observed. Flagged outlier sequences were removed by the Ape R package v5.7-1 (Popescu et al., 2012) and the regenerated phylogenies visualised by the ggtree R package v3.6.2 (Yu, 2022). Ancestral state reconstruction to estimate the imports and export events into and from Kilifi and Uganda locations was undertaken using the mugration TreeTime package (Sagulenko et al., 2017). The number of state changes was determined using a custom python script (https://github.com/CERI-KRISP/SARS_CoV_2_VOC_dissemination) and plotted using the ggplot2 package v3.4.3 in R.

## Results

### IBV detections in Kenya (Kilifi) and Uganda

A total of 18,037 samples were tested for IBV in the KCH, KHDSS and COAST studies whereby 173 (1.0%) tested positive. The number of IBV cases were variable from month to month in the years having detections (**Figure 2a**). Every year of the study samples sequenced, except 2018 and 2020 for Uganda and 2018 for Kenya. Sequencing was attempted for 151 samples (87.0%) that had sufficient sample volume available (>140 μl) and these yielded 83 (55.0%) complete or near complete genomes (>70.0% coverage). Of these, 7 (8.0%) clustered with B/Yamagata lineage reference strains and 76 (91.0%) clustered with B/Victoria lineage reference strains (**Figure 2b**).

**Figure 2.**
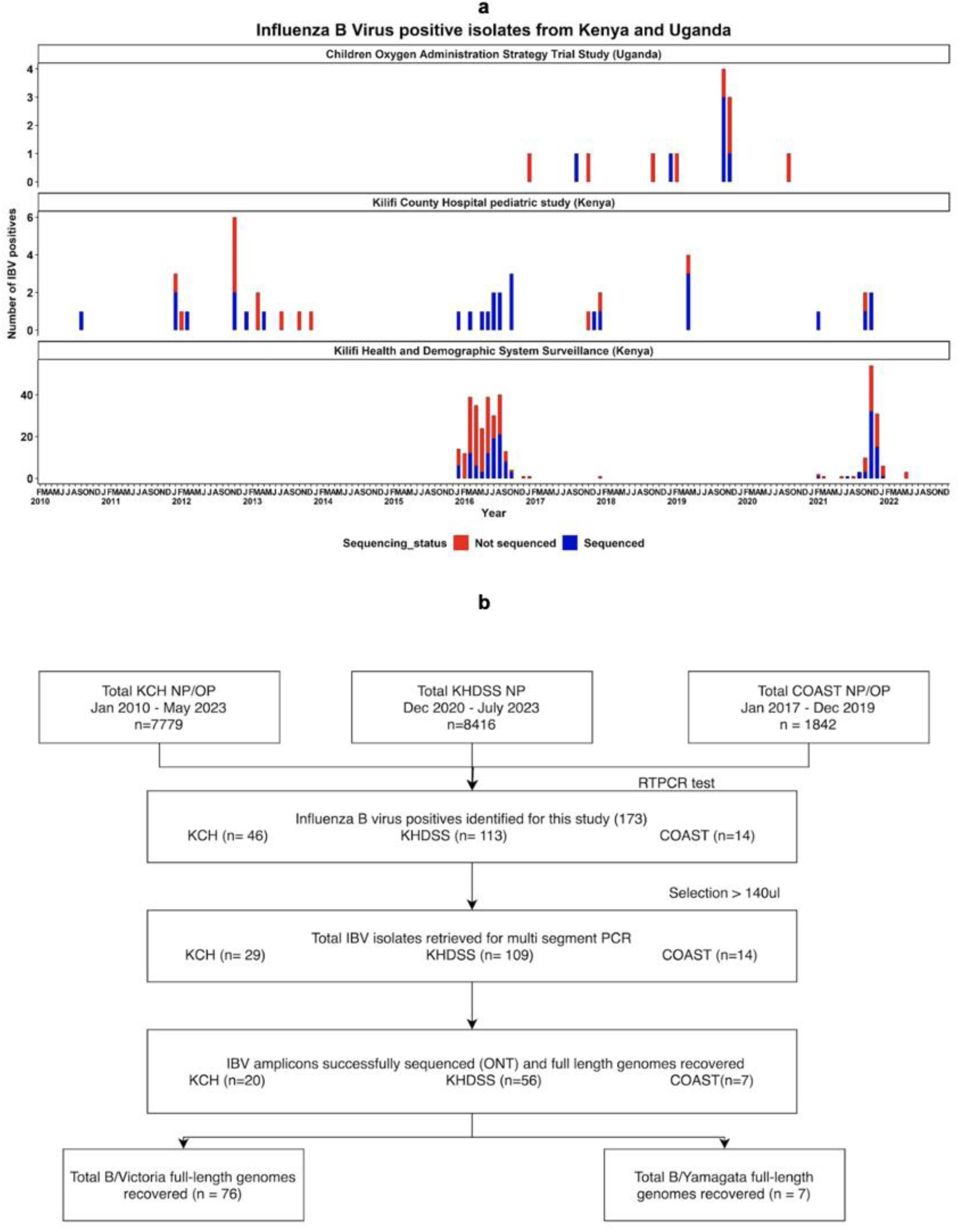
Source, temporal distribution and sample laboratory processing of IBV positives detected in Kilifi and Uganda during the KHDSS, KCH and COAST surveillance period. **a,** Bargraph showing the number of IBV detections per study per month across the surveillance period. Bars are coloured by the proportions of isolates sequenced or not sequenced. **b,** Sample flow chart showing the number of samples processed and whole genome sequencing yield from the three studies.

### IBV lineages, clades and subclades circulating in Kilifi and Uganda

IBV are classified into lineages and subsequent clades and subclades by the characteristic amino acid changes on the HA segment (**Figure 3a**). Sequencing yielded near complete genomes from 3 years in Uganda and from 9 years in Kilifi (**Figure 3b**). Notably, B/Victoria lineage predominated in 6 years in Kilifi and in all 3 surveillance years in Uganda. Co-circulation with B/Yamagata strains was observed in 2016 in Kilifi and in 2019 in Uganda. Lineage predominance in Kilifi and Uganda was consistent with the global trends inferred from the complete global dataset from the GISAID database (**Supplementary figure 1**) except for the 2017 season in Uganda where only one sample was available. The lack of detected B/Yamagata strains among Kilifi and Uganda strains after 2019 is consistent with WHO and global reports of no confirmed B/Yamagata cases among samples collected from March 2020 onwards (WHO, 2023).

**Figure 3.**
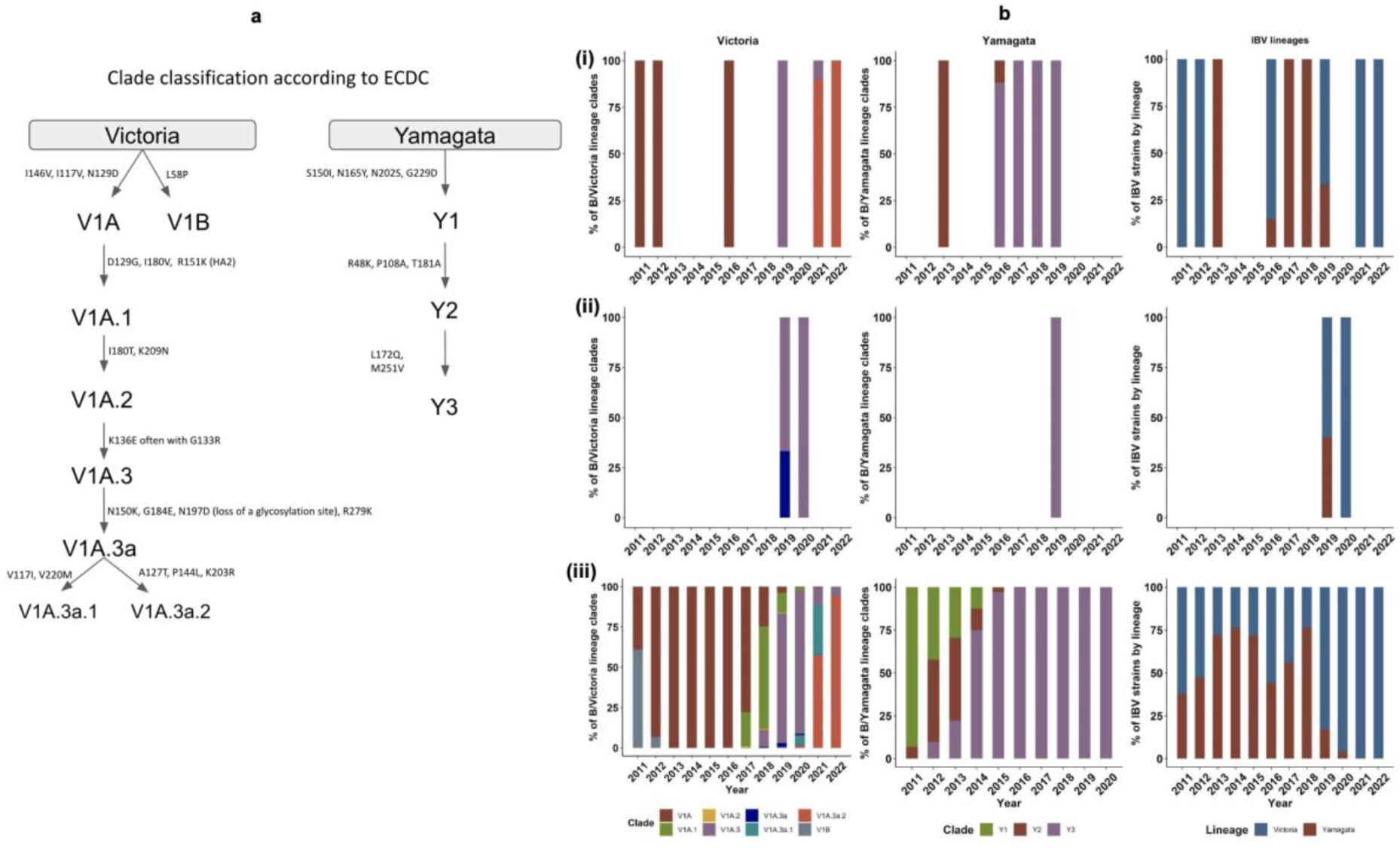
Identification and temporal distribution of circulating lineage, clade and subclades in Kilifi and Uganda. **a**, Cartoon representation of IBV clade and subclade classification according to ECDC annotated with clade defining mutations. **b**, Temporal patterns of IBV lineages, clades and subclades among subtyped sequences observed in Kilifi and Uganda during the surveillance period in comparison with global trends. Lineages, clades and subclades were assigned according to ECDC guidelines (**Table 1**). Panels **(i)**, **(ii)**, **(iii)** represent clade and subclade distribution of B/Victoria strains (left), B/Yamagata strains (centre) and lineage distribution (right) of Kilifi, Uganda and global strains respectively.

The B/Victoria lineage strains in Kilifi were classified into clade V1A and further into subclade V1A.3 and V1A.3a.2 while Uganda strains clustered with subclade V1A.3 and its subclade V1A.3a reference strains (**Supplementary figure 2**). Notably, there was an observed clade shift to the V1A.3a.2 subclade as from 2020 which was one of the peak years of the COVID-19 pandemic characterised by low influenza detections, with the clade being predominant globally to date (May 2023) (**Figure 3b**). B/Yamagata lineage strains circulating in Kilifi fell into clades 2 and 3 whereas only clade 3 viruses were detected in Uganda. As observed globally, clade 2 strains in Kilifi were observed until 2016 when clade 3 strains became predominant (**Figure 3b**).

### Phylogenetic analysis of IBV gene segments

The eight IBV gene segments from the Kilifi and Uganda strains were analysed for genomic clustering with known clade and subclade representative viruses (**Table 1**). Consistent phylogenetic clustering patterns was observed all segments with no reassortants detected (**Supplementary figure 3**). Five distinct phylogenetic clusters, defined by clustering of strains with clade and subclade reference strains with high bootstrap support (> 70.0%), were identified in the Kilifi strains and three observed in Uganda strains for both lineages (**Supplementary figure 2**).

B/Yamagata clade 2 strains clustered with the B/Massachusetts/02/2021 strain bearing the clade defining mutations. Clade 3 strains were B/Phukhet/3073/2013-like with additional N116K, K298E and E312K mutations previously detected in this group (ECDC 2023). The B/Victoria strains from 2011 and 2012 grouped with the B/Brisbane/60/2008 reference strain with an additional I146V mutation (supplementary file 2). Kilifi and Uganda B/Victoria lineage strains from 2019 to 2022 were deletion variants bearing the 3-aa deletion 162-164 in the HA segment. The clade 1A.3 strains clustered with the B/Washington/02/2019 reference strain but were grouped into strains containing the subclade defining mutations (**Table 1**) which comprised three Ugandan strains and one Kilifi strain and a subcluster comprising six Kilifi strains with K75E, E128K, T155A, G230N mutations which have been previously detected in Netherlands and other Kenyan strains (ECDC 2023; ECDC 2021). The subclade 1A.3a split into the 1A.3a.1 and 1A.3a.2 subclades with Kilifi strains from the latter clustering with the B/Austria/1359417/2021 having subclade defining mutations. In addition, a subcluster with T182A, T221A, N126S and S286F mutations within the subclade were identified (supplementary file 2).

### Glycosylation analysis of the HA segment

A total of eleven N-linked glycosylation potential sites were identified in the B/Victoria strains, seven on the HA1 domain at positions 25, 59, 145, 166, 233, 304 and 333 and four in the HA2 domain at positions 507, 533, 546 and 578 (**Supplementary figure 4)**. A loss in the glycosylation site at position 197 of the HA1 domain was observed in subclade 1A.3a and 1A.3a.2 strains (**Supplementary figure 4**). Gain or loss at this site affects the receptor-binding properties and modulates the antigenicity of the virus (Saito et al., 2004). Kilifi and Uganda B/Yamagata strains possessed 7 potential N-linked glycosylation sites in the HA1 domain at positions 25, 59, 145, 167, 196, 303 and 332 and at positions 506, 532, 545 and 577 in the HA2 domain (**Supplementary figure 4**).

### Antigenic mutations along the HA backbone

IBV contains four main antigenic epitopes located in the HA1 domain comprising the 120 loop (116 - 137), 150 loop (141-150), 160 loop (162-167) and the 190 helix (194-202) (Wang et al., 2008). Antigenic mutations in the B/Victoria strains majorly accumulated in the 120 and 190 loops (**Figure 4**). Comparison with global strains identified two unique amino acid changes in V1A clade (T147A, H27Y), two in the B/Yamagata clade 2 viruses (K129N, S285A) and one (N164Y) in clade 3 strains. However, these unique changes were observed in less than three local strains. A majority of epitope and non-epitope amino acid changes detected locally in both lineages were also detected in global strains, suggesting similar clade and subclade antigenic profiles.

**Figure 4.**
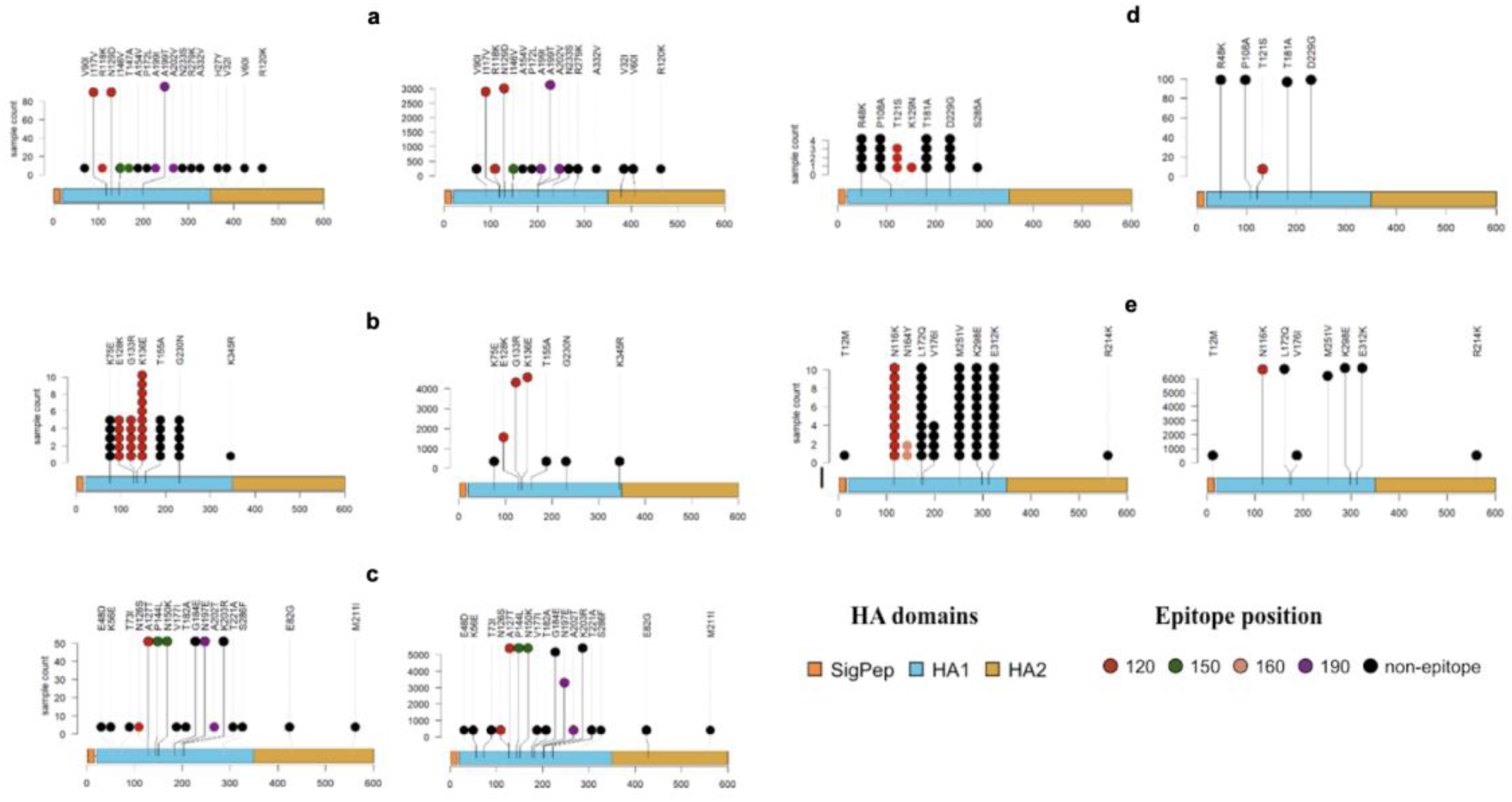
Distribution of amino acid changes along the IBV HA segment of Kilifi and Uganda strains in comparison with global strains. Lolliplot pairs labelled **a-e** represent the amino acid changes identified in the V1A, V1A.3, V1A.3a.2, Y2 and Y3 clades respectively. Lolliplots on the left side of each pair show the amino acid changes identified along the HA segment of Kilifi and Uganda strains whereas lolliplots on the right represent the number of amino acid changes also identified among the global strains. Epitope and non-epitope amino acid changes are colour coded (see key). The height of the plot represents the number of samples containing the amino acid change. Lolliplot branches in grey represent amino acid changes identified in three sequences or less. The plots were generated using the Trackviewer package in R.

### Global context of Kenya and Uganda IBV strains

The global phylogenetic context of IBV lineages was examined using an approximate maximum likelihood (ML) approach in TreeTime (Sagulenko et al., 2017). The global dataset included 2994 B/Victoria and 2088 B/Yamagata strains subsampled from six continents between 2010 and 2022. For both lineages, Kilifi (Kenya) and Uganda strains clustered with strains from both Africa and the rest of the world (**Figure 5**).

**Figure 5.**
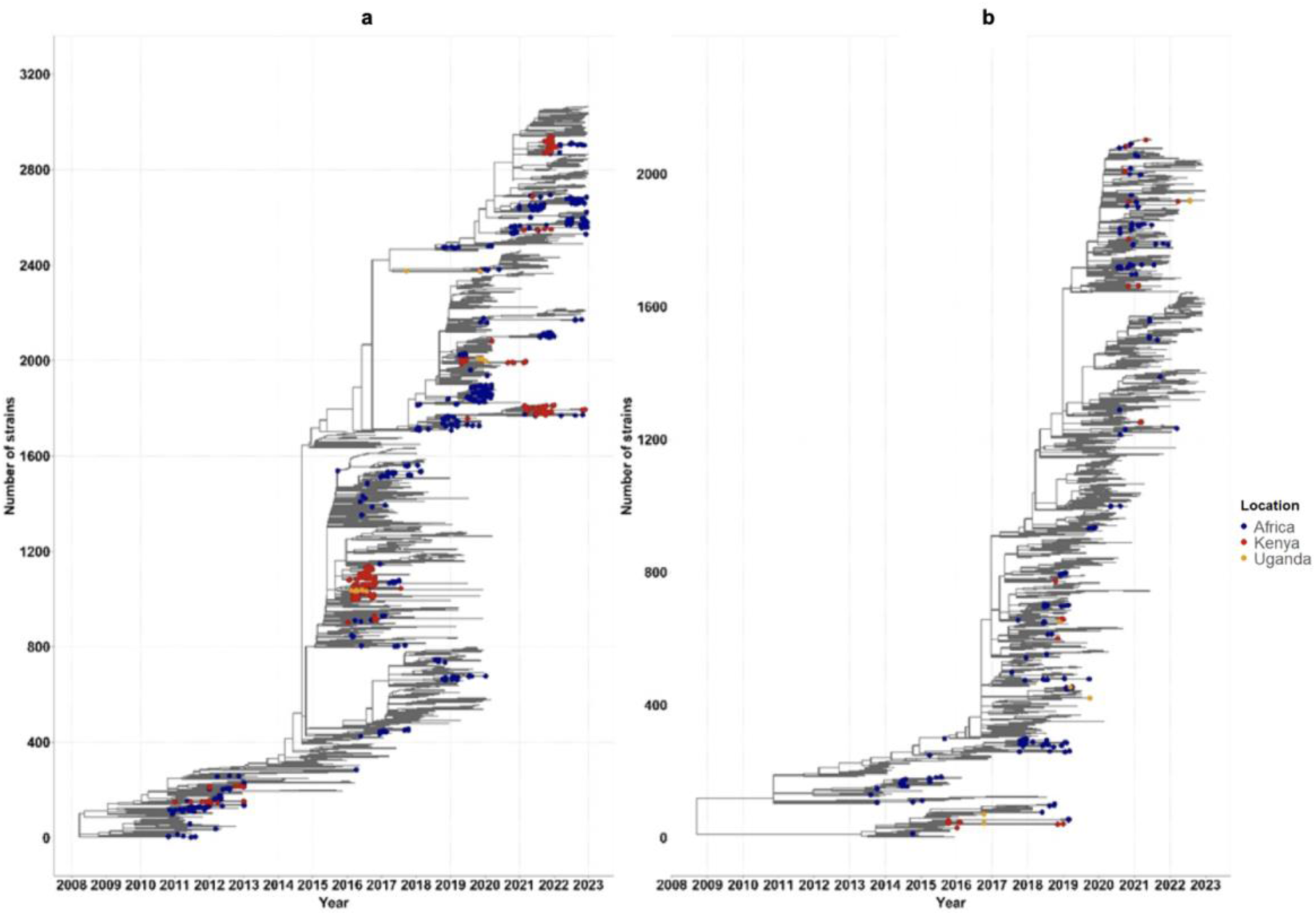
Global phylogenetic context of Kenya, Uganda and global IBV strains. **a, b** represent time-resolved phylogenies of global, Kilifi and Uganda IBV strains of B/Victoria and B/Yamagata lineages respectively inferred using TreeTime. Unlabelled tips represent strains from other continents.

Multiple distinct clusters of Kilifi and Uganda strains were identified across the years (**Figure 5**), suggesting multiple IBV introductions into the countries followed by onward local transmission. Ancestral state reconstruction of these phylogenies identified 7 exports, 28 import and 68 inter-facility events among Kilifi B/Victoria strains while Uganda strains exhibited 2 inter-facility events and 3 imports, 2 originating from Kenya (supplementary file 5). B/Yamagata Kilifi strains had 8 imports and 2 inter-facility events while 2 import events were detected in Uganda strains (**Supplementary figure 5**).

## Discussion

IBV epidemiology and evolution in East Africa is understudied, despite its significant disease burden in the region (Sambala et al., 2018; Seleka et al., 2017; Langat et al., 2017; Nyasimi et al., 2020; Touré et al., 2022). We analysed circulating IBV strains over nine years in Kilifi, Kenya, and four years in Uganda. Both IBV lineages were detected with alternating lineage predominance and co-circulation in some years. The lineage and clade predominance patterns were highly consistent with global trends. No B/Yamagata strains were detected from 2019 onward, contrary to B/Victoria strains. A clade shift within B/Victoria lineage to subclade V1A.3a.2 was observed in 2021 in Kilifi. The IBV strains detected were non-reassortants having additional amino acid changes and showing a loss in a potential N-glycosylation site.

Lineage patterns in Kilifi between 2012 and 2016 mirrored those observed in a previous study in Kenya (Emukule, Otiato, et al., 2019). The B/Victoria lineage predominated most years in both Kenya and Uganda compared to the B/Yamagata lineage. However, the clade dominance was less predictable, and infections appeared to be sporadic, punctuated with periods of more intense transmission. This complicates the optimal selection of vaccine strains. No B/Yamagata strains were detected since 2019, and this is consistent with global reports suggesting the possible extinction of the lineage (Paget et al., 2022; Koutsakos et al., 2021). Although the extinction of other B/Yamagata clades such as the B/Yamanashi/166/98 which was in circulation until 2002, has been documented (Langat et al., 2017), the potential resurgence of seemingly extinct influenza virus lineages remains a concern in the future, as was the case in the reemergence of A (H1N1) in 1977 (Rozo & Gronvall, 2015).

Amino acid changes in IBV HA and NA surface proteins guide the selection of suitable vaccine strains. The detection of subclades whose additional amino acid changes were first detected in Kilifi strains and subsequently identified in the Netherlands highlights the importance of global IBV genomic surveillance for vaccine strain reformulation. The highly conserved NA gene encodes a sialidase enzyme that catalyses virion release from the host cell and has been a target for NA inhibitors (McAuley et al., 2019; Holmes et al., 2021). Over time, mutations conferring resistance to these inhibitors have been identified (Sheu et al., 2008) however, these were not identified in the Kilifi and Uganda strains. The absence of oseltamivir-zanamivir resistance amino acid changes in Kilifi and Uganda strains indicates the effectiveness of these interventions and might be due to the infrequent use of these drugs in local communities.

N-linked glycosylation plays a crucial role in protein folding, pH stability and modulation of virus antigenicity in the HA stalk and near RBS regions (York et al., 2019). The loss of the 196/197 glycosylation site observed here has previously been attributed to egg adaptation and has an effect on binding to α-2,3-linked sialic acid receptor, hence affecting virus antigenicity (Chen et al., 2008).

This study has several limitations. First, our sample size is small creating significant uncertainty in some of our inferences. There were some years with low number or no IBV positive isolates to sequence. The parent study periods only partially overlapped, with the surveillance in Uganda happening in few seasons. Thus, the long-term transmission patterns of the virus in Kenya and Uganda could not be concluded. The Kenya surveillance only sampled one of the 47 counties in the country. Second, the limited number of positive IBV isolates from Uganda impacts the generalizability of lineage predominance, intra-and inter-country viral flow within a season.

In conclusion, IBV lineages in East Africa circulate with an alternating predominance, with a progressive turnover of clades undergoing continuous evolution through antigenic drift and glycosylation mechanisms. The observed congruence with global patterns shows that these strains are quite like global strains, hence suitability of global interventions such as vaccines for local use.

## Data Availability

All data produced are available online in Gen Bank under accession numbers OR240875-OR240981 and OR267522-OR268202

https://www.ncbi.nlm.nih.gov/nuccore/?term=OR267522:OR268202[accn]

https://www.ncbi.nlm.nih.gov/nuccore/?term=OR240875:OR240981[accn]

## Acknowledgments

We thank all the participants and staff from all the centres participating in the COAST trial. We thank the Trial Steering Committee for its support of the trial and members of the Pathogen Epidemiology and Omics (PEO) Group at KWTRP for their support in sample processing and analysing the reported results. We acknowledge all the authors and laboratories that uploaded their datasets in the GISAIDTM EPIFLU database where we sourced global data.

## Data availability

High throughput sequence datasets for IBV segments are available in GenBank under accession numbers OR240875-OR240981 and OR267522-OR268202.

## Code availability

All the scripts used for analysis in this study are available at the Github repository: https://github.com/Kiage24/Influenza_B_transmission_dynamics

## Author contribution statement

**Brenda K Nyarango:** Conceptualization and design, methodology, laboratory and bioinformatics data analysis, visualization, drafting and review of article

**D. Collins Owuor, Everlyne M. Isoe and George Githinji**: Conceptualization and design, bioinformatics analysis, supervision, review of article

**Joyce Nyiro, Sarah Kiguli, Peter Olupot-Olupot**: Field data collection, review of article

**Esther Katama**: Data management; data retrieval and archiving

**Martin Mutunga & Robinson Cheruiyot**: Laboratory analysis-viral genome purifications and amplification

**Timothy Makori**: Laboratory analysis-viral sequencing and interpretation of laboratory data

**Arnold Lambisia**: Bioinformatics analysis and interpretation of data, review of article **Kathryn Maitland**: Funding acquisition, conceptualization and design, data acquisition, supervision, review of article

**Charles N Agoti**: Funding acquisition, conceptualization and design, data acquisition, supervision, drafting and review of article

**Conflict of interest**: None declared

