## Supplementary figures for "Epidemiological dynamics of Influenza B virus across multiple seasons in Kenya and Uganda inferred from sequence data, 2010-2021": Supplementary_figure_5.pdf (from another version of this preprint: version 1, 2025-03-07)

a). Import and export events among Kenya and Uganda B/Victoria IBV strains

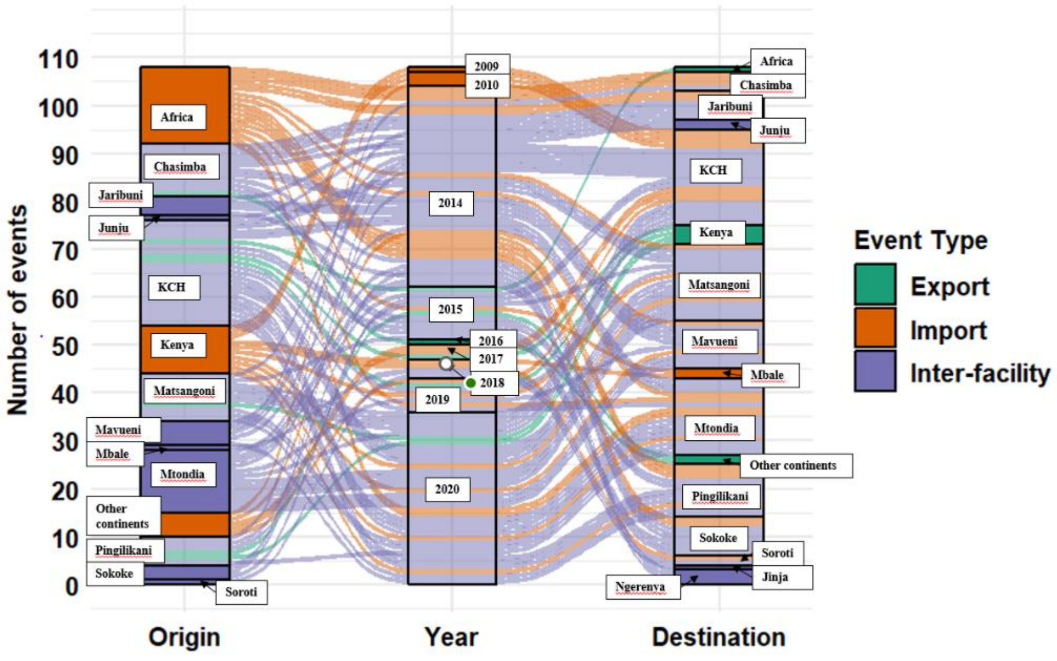


b). Import and export events among Kenya and Uganda B/Yamagata IBV strains

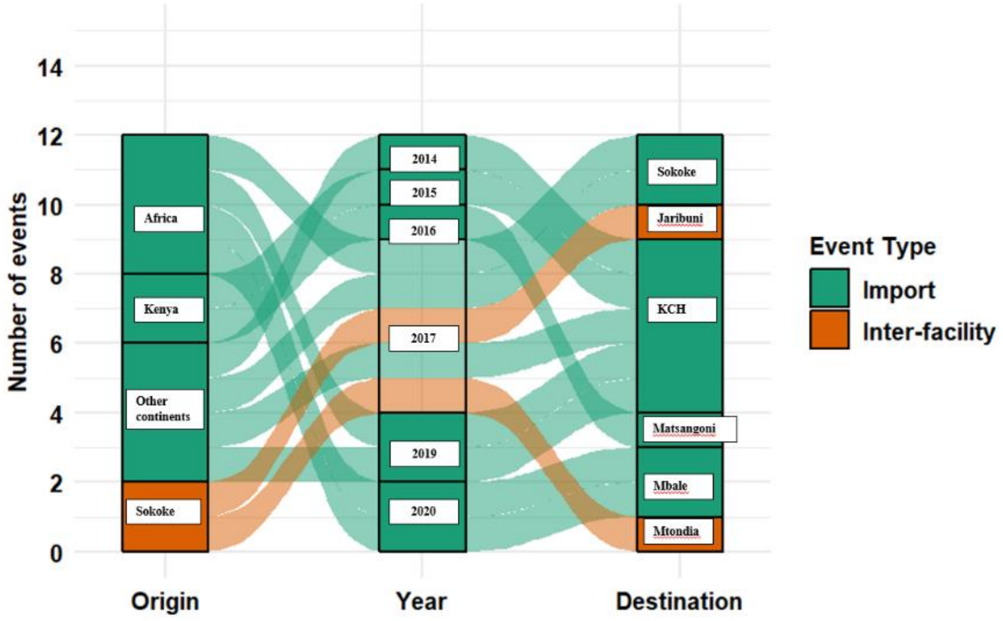

(a) and (b) are alluvial plots showing the origin, destination and year of virus import and export events of the B/Victoria and B/Yamagata strains respectively. The plots are coloured according to the type of event identified (see legend).
