## Supplementary figures for "Epidemiological dynamics of Influenza B virus across multiple seasons in Kenya and Uganda inferred from sequence data, 2010-2021": Suppplementary_figure_1.pdf (from another version of this preprint: version 1, 2025-03-07)

**Flowgram showing the cleaning and random subsampling criteria for GISAID global contamporaneous sequences.**

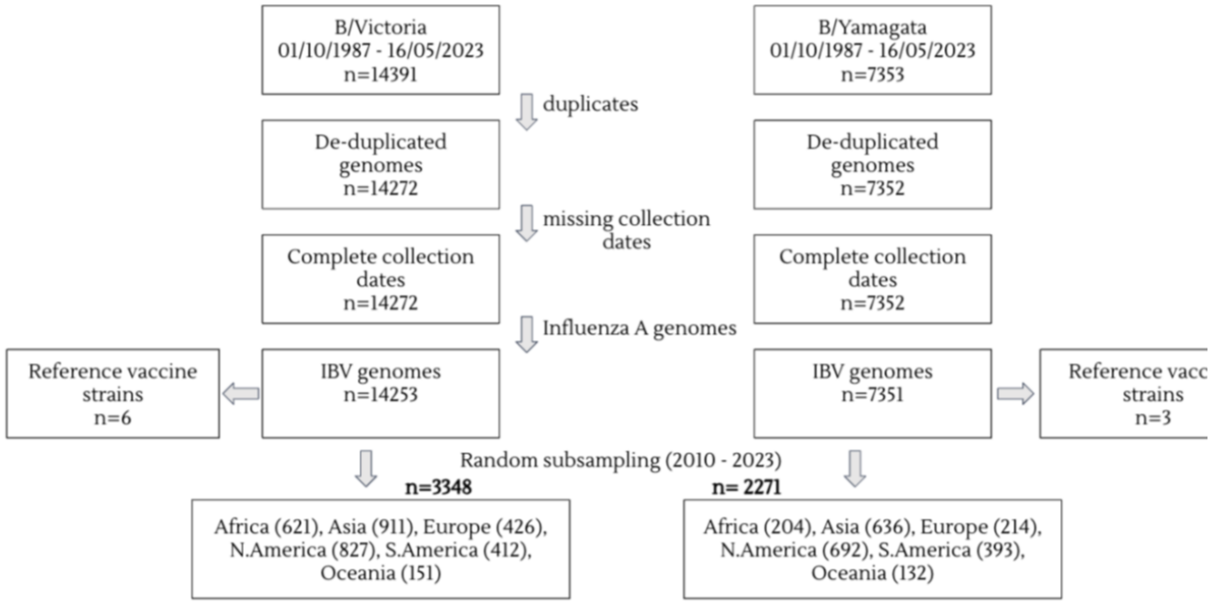
