## Supplementary figures for "Epidemiological dynamics of Influenza B virus across multiple seasons in Kenya and Uganda inferred from sequence data, 2010-2021": Suppplementary_figure_2.pdf (from another version of this preprint: version 1, 2025-03-07)

**Phylogenetic lineage, clade and subclade classification of IBV in Kilifi and Uganda based on the HA segment.**

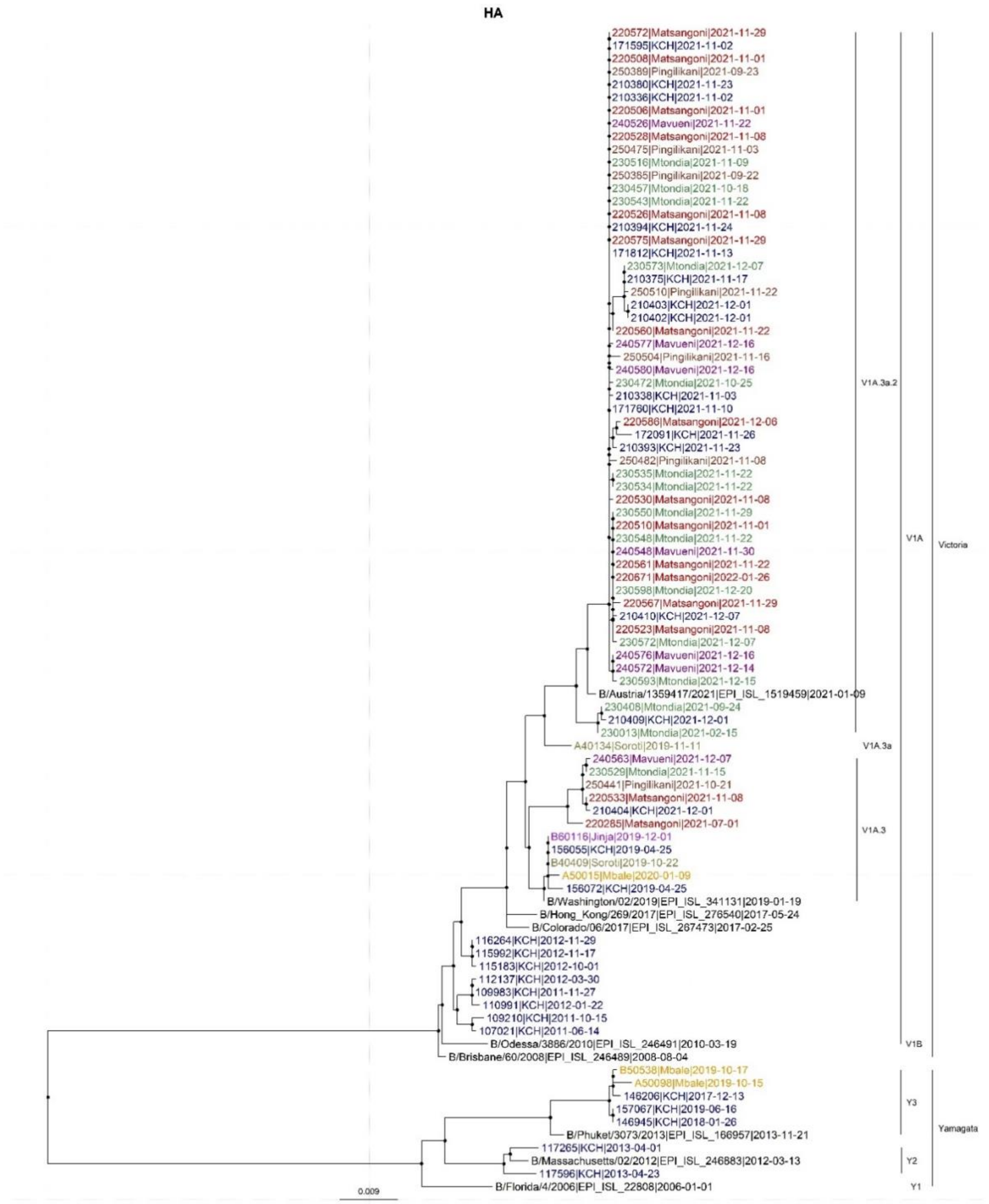


*Branches are annotated with amino acid alterations identified along the HA trunk and branch tip labels are coloured according to sample locations with the exception of reference strains represented in black (see legend). Amino acid changes defining*

*clades and subclades are in bold. Nodes having >70% bootstrap reliability are marked with black shaded circular nodes.*
