## Supplementary figures for "Epidemiological dynamics of Influenza B virus across multiple seasons in Kenya and Uganda inferred from sequence data, 2010-2021": Suppplementary_figure_3.pdf (from another version of this preprint: version 1, 2025-03-07)

**a). Genetic relatedness of Kilifi and Uganda IBV strains.**

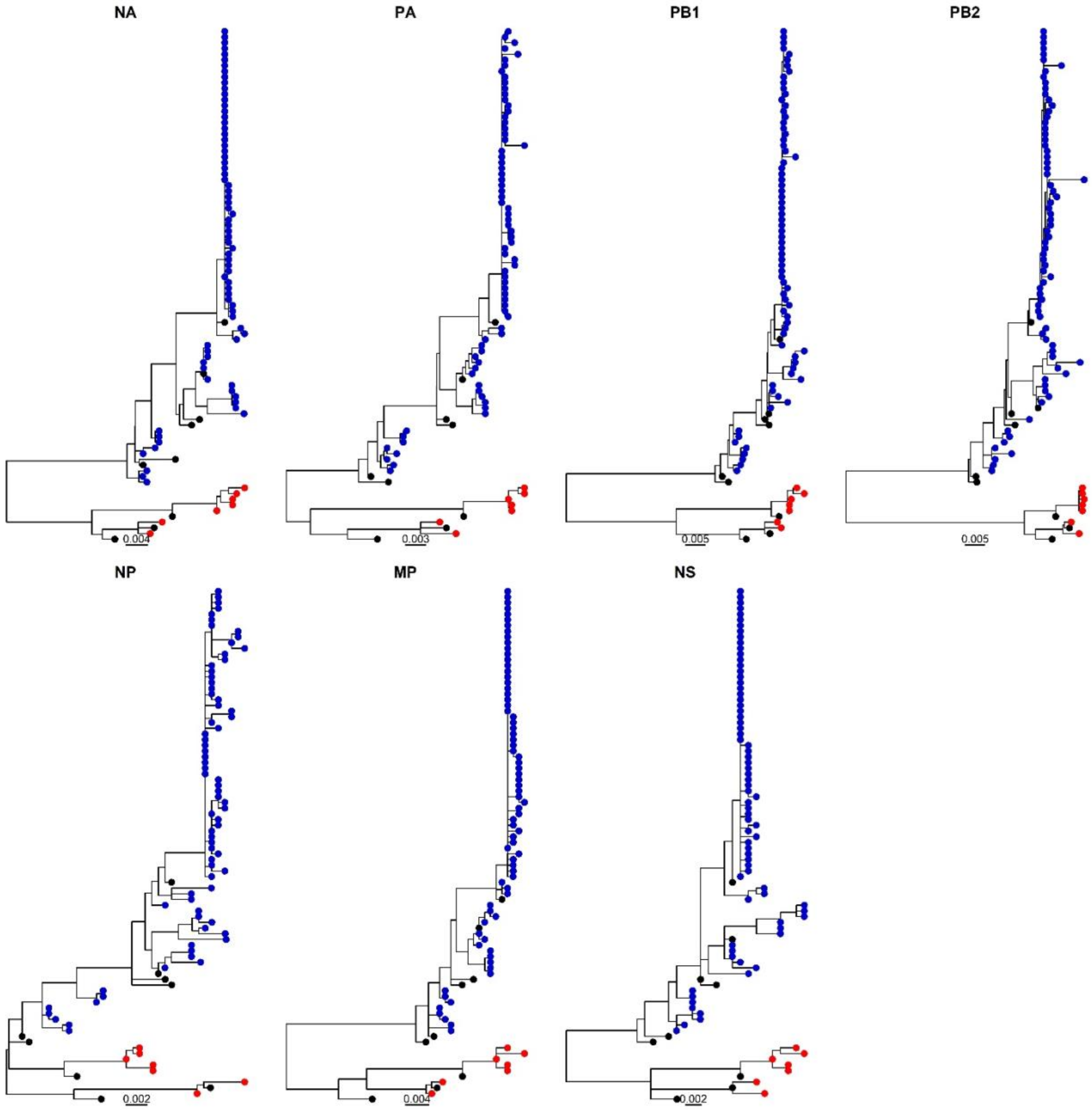


*Per segment maximum likelihood phylogenies were inferred from 91 IBV strains from Kilifi (75), Uganda (6) and 10 reference strains.*

**b). Tanglegrams showing the phylogenetic congruency between IBV gene segments.**

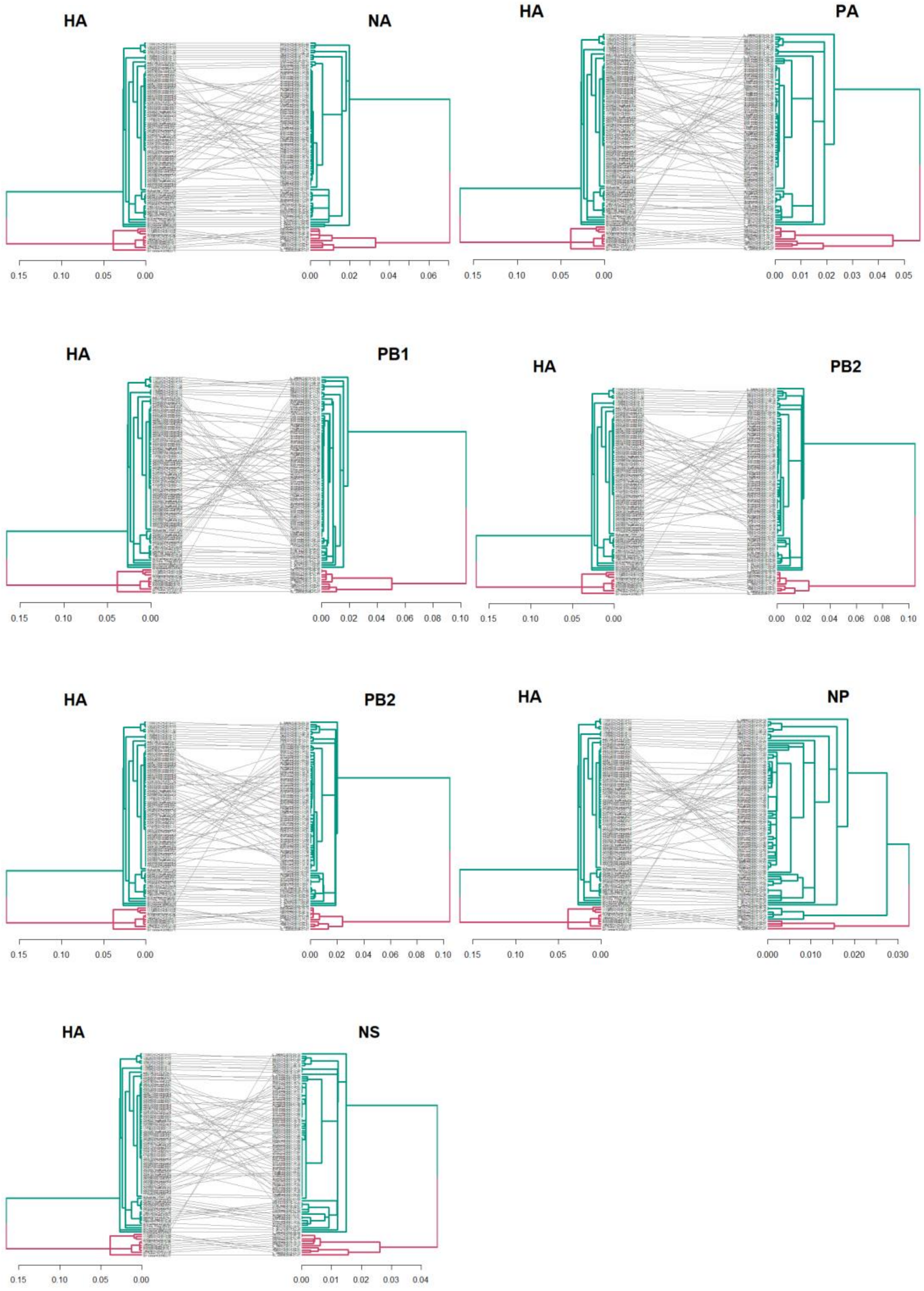

*B/Yamagata strains are represented by the red branches while B/Victoria strains are represented by the green branches.*
