## Supplementary figures for "Epidemiological dynamics of Influenza B virus across multiple seasons in Kenya and Uganda inferred from sequence data, 2010-2021": Suppplementary_figure_4.pdf (from another version of this preprint: version 1, 2025-03-07)

**Bar plots showing N-glycosylation site proportions in B/Victoria and B/Yamagata strains from Kilifi, Kenya and Uganda.**

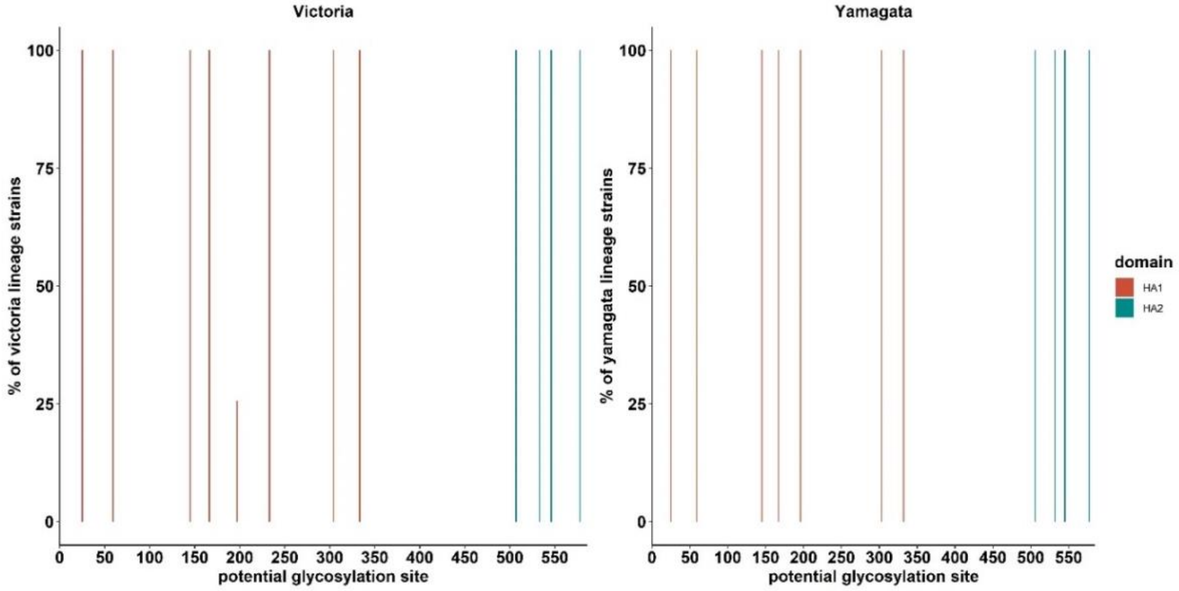
